# Genetic confounding in the associations between maternal health and autism

**DOI:** 10.64898/2026.04.16.26351033

**Authors:** Elias Speleman Arildskov, Viktor H. Ahlqvist, Vahe Khachadourian, Zeynep Asgel, Diana Schendel, Stefan Nygaard Hansen, Jakob Grove, Magdalena Janecka

## Abstract

The etiology of autism is influenced by genetic and non-genetic factors, with observational studies suggesting associations between early maternal health diagnoses and offspring autism. However, these associations may partly reflect shared familial genetic liability rather than direct causal effects.

Using comprehensive national health registers and individual-level genetic data from the iPSYCH cohort (N=117,542), we examined whether maternal health diagnoses are associated with offspring polygenic scores (PGS) for autism. Such associations between maternal health and offspring autism would indicate shared genetic factors and the possibility of genetic confounding in the observational associations. We also tested such associations with PGSs for other neuropsychiatric and neurodevelopmental conditions that are genetically correlated with autism, but with better-powered PGS (due to larger GWAS sample sizes and likely more polygenic genetic architecture), as well as height, a negative control.

Several maternal diagnoses were nominally associated with autism PGS in the child, including, e.g., certain obstetric complications, asthma, and obesity. After adjustment for multiple testing, the only statistically significant results included those between maternal diagnoses, predominantly psychiatric, and other neuropsychiatric and neurodevelopmental PGSs in the child. Sensitivity analyses confirmed the robustness of our results across exposure windows, diagnostic settings, and socioeconomic adjustments.

These findings indicate that maternal diagnoses associated with autism partially reflect shared genetic liabilities between mothers and their children. However, such genetic effects, as captured by child PGS do not fully explain the observed associations, suggesting additional factors, including e.g., non-genetic familial factors, rare variants, and indirect effects.

## Introduction

Autism is a neurodevelopmental condition, typically diagnosed in childhood based on behavioral and developmental observations^1^. The etiology of autism is complex, with high familial heritability^2,3^, the role of spontaneous, *de novo* genetic mutations in a subset of diagnosed individuals^4^, and the putative contribution of environmental factors^5^.

While multiple environmental factors, particularly those occurring in prenatal or early postnatal stages, have been implicated in the etiology of autism^6–8^, the causal evidence for their role remains inconclusive. Our^9^ and others’^10–12^ work has indicated that at least some of the observational associations between early-life exposures and autism likely arise due to familial confounding, *i*.*e*., family-level factors like socioeconomic status, pollution, or genetic variation, that are associated with the likelihood of both the perinatal exposure and autism. While these studies challenge the role of a causal contribution of at least some of the early-life exposures to autism, they stop short of providing a mechanistic understanding of autism.

The familial confounding factors could represent a wide range of genetic and non-genetic influences relevant to autism etiology. Additionally, in line with the liability threshold model^13^ of a multi-factorial condition like autism, many such familial, causal factors may act in an additive manner, with none of them essential for autism development. For example, while genetic factors are likely one of the key causal factors in autism etiology^2,4^, they have a weaker association with autism in individuals with a history of certain obstetric complications^14,15^. Therefore, beyond identifying a single source of familial confounding, research still needs to highlight how different familial factors come together to contribute to complex and heterogeneous etiology of autism.

In the current project, we focused specifically on the role of familial *genetic* factors and the extent to which they contribute to the observational associations between maternal health in pregnancy and autism^9^. Evidence for an enrichment of genetic variants associated with autism in children of females with specific diagnoses (*e*.*g*., asthma, depression), would entail a possibility of genetic confounding in the previously reported observational associations between maternal health and autism^9,16,17^. Genetic variants could contribute to such confounding via their (i) direct, pleiotropic effects on both maternal and child conditions, impacting child’s neurodevelopment to the extent they are transmitted from mother to child^11,12^; (ii) indirect, pleiotropic effects, affecting the risk of the maternal condition and the in utero environment — thus operating irrespective of the variant transmission to the child^18^; or, (iii) without pleiotropy, if genetic variation in the family impacts the likelihood of the maternal and child diagnoses, e.g., through its effects on the family’s access to healthcare.

To explore the potential genetic sources of familial confounding in the associations between maternal health and autism, we used the large-scale iPSYCH resource^19,20^ with individual-level genetic, medical, and socioeconomic data on over 134,000 individuals in Denmark. This rich dataset, complete with multi-generational pedigree data and known sampling weights, enabled us to investigate the link between maternal health status and child’s genetic liability to autism and related neuropsychiatric conditions, derive population-based estimates of such associations, and implement a series of sensitivity analyses to assess the robustness of our modelling.

## Methods

### Study population

The source population for the iPSYCH study^19^ included all people born in Denmark between May 1^st^ 1981 and December 31^st^ 2008, with a known mother, who were alive, and residing in Denmark at the age of 1. From those, all individuals diagnosed with one or more of the following conditions by December 31^st^ 2015 (December 31^st^ 2016 for anorexia) were selected into iPSYCH: autism, attention-deficit/hyperactivity disorder (ADHD), schizophrenia or schizophrenia spectrum disorder, affective disorder, postpartum depression, anorexia, and bipolar disorder (see **Table S1** for the ICD codes). A random sample from the source population was also selected (subcohort, N = 50,615; note that, due to random sampling, the subcohort included individuals with mental health diagnoses, at the population prevalence level). From the iPSYCH participants, those without any of the exclusion criteria listed below were included in the current study.

Exclusion criteria in this study included (a) inclusion in iPSYCH based on a temporary diagnosis which was later removed; (b) missing genotype data (due to the neonatal blood spot not found in the biobank, insufficiency of the material, or genotyping failure); (c) discrepancy between the ID of the legal mother in the Central Person Register and the maternal ID identified in the Medical Birth Registry, or the lack of child-mother dyad linkage in the Medical Birth Registry; (d) missing covariate data. Additionally, (e) amongst pairs of individuals with a high degree of relatedness 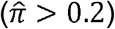, one individual was removed from each pair; where possible, within each pair we removed the individual who was related to most people in the sample, or not a member of the subcohort, otherwise one individual from each pair was removed at random; and (f) individuals of non-European ancestry were not included in the main analyses, to ensure that our results are not driven by between-ancestry, artefactual differences in the PGS and potential differences in the prevalence of certain maternal conditions. Non-European ancestry individuals were identified based on an ellipsoid using the first 3 principal components (PCs), centered and scaled around the mean and 8 standard deviations of those whose parents and grandparents are all known to have been born in Denmark (3^rd^ generation Danish born); NB. these individuals were included in one of the sensitivity analyses. A flowchart with the number of individuals excluded based on each criterion is presented in **Figure S1**.

### Exposure

The exposures were the maternal diagnoses associated with autism in a prior, diagnosis-wide, population-based study in Denmark^9^ comprised of mostly maternal psychiatric, cardiometabolic, and obstetric diagnoses identified through the National Patient Register or the Psychiatric Central Research Register using level-3 ICD-10 diagnoses (e.g., *E66: Overweight and obesity;* **Table S2a**). For completion, we also included maternal diagnoses associated with autism in other studies, including rheumatoid arthritis^1617^, inflammatory bowel disease^17^, intrahepatic cholestasis of pregnancy^21^, and maternal cytomegalovirus infection^22^ (**Table S2b**). To match ICD-8 and ICD-10 codes and ascertain the diagnoses across the entire study period, we used the conversion table compiled for use within Danish registers^23^.

We ascertained maternal *lifetime* diagnoses, reflecting our focus on genetic liabilities associated with maternal conditions, rather than their acute consequences in pregnancy. Individuals with ICD8 diagnosis code modifiers indicating uncertainty (suspected, but without sufficient evidence, or those that were not found) were considered unexposed, unless there was another record of the diagnosis without the uncertainty code (a similar diagnostic code modifier does not exist for ICD-10).

Given the temporal changes in the diagnostic criteria and recording (transition from ICD-8 to ICD-10 in 1994, and nationwide inclusion of outpatient diagnoses in the Danish registers only since 1995), we conducted a series of sensitivity analyses to ensure lifetime ascertainment of exposure did not introduce a systematic bias based on maternal or child’s birth year.

### Outcome

The main outcome was the index child’s polygenic score for autism (ASD PGS), calculated using BOLT-LMM and LDpred2 software^24^ to obtain a meta-PGS^25^. This process followed a 5-fold cross-validation scheme to avoid overfitting, utilizing 80% of the data for training, 10% for validation, and 10% for testing. The weights were generated in the validation set and standardized based on the unweighted mean and standard deviation in the sub-cohort.

To derive the meta-PGS, first, an external PGS (*PGS*_*ext*_) was calculated with LDpred2 using the GWAS summary statistics^26^, excluding the iPSYCH samples. Then an internal PGS (*PGS*_*int*_) was calculated based on per-SNP prediction betas found using BOLT-LMM in the iPSYCH sample. Lastly, the meta-PGS was constructed, using the linear combination of PGS_ext_ and PGS_int_, with weights *w*_*int*_ and *w*_*ext*_ (*metaPGS= w*_*0*_*+ w*_*int*_*PGS*_*int*_ *+ w*_*ext*_*PGS*_*ext*_).

We conducted additional analyses where outcomes were the PGSs of other neurodevelopmental and psychiatric diagnoses with high genetic correlation with autism^27^, including: major depressive disorder (MDD)^28^; attention-deficit/hyperactivity disorder (ADHD)^29^, schizophrenia (SCZ)^30^ and neuroticism (NEUR)^31^. Additionally, we included height PGS^32^ as a negative control. The PGSs for MDD, ADHD and SCZ were calculated as described for ASD PGS; height and neuroticism PGS were derived using LDpred2-auto^24^.

### Covariates

In the main model, we accounted for the top 5 genetic principal components (PCs), calculated using the smart PCA method^33^ and *smartsnp* R package^34^. We also adjusted for the genotyping chip due to potential systematic variation in PGS based on the chip used, and child’s legal sex and year of birth, in order to account for the differences in the prevalence of different iPSYCH conditions across the study birth years and by sex.

In additional analyses, along with the covariates in the main model, we included factors with a potential impact on maternal likelihood of receiving certain diagnoses, including: maternal age at child’s birth; healthcare utilization (defined as the number of distinct days the mother had an ICD-8 or ICD-10 diagnosis in each year with available maternal diagnostic data); maternal highest education attained up to the year the child was born, or, if missing, highest attained in any of the previous years for which the data were available; maternal income the year before the birth of the child, ensuring the variable reflects income from employment, rather than parental leave pay (to mitigate against potential outliers and ensure comparability between birth years, this variable was calculated as the rank of the index person’s maternal income for the given year, divided by the number of participant observations in the given year (bound between 0 and 1)). All continuous variables, aside from the PCs, are included using restricted cubic splines with 4 knots placed at percentiles 5, 35, 65 and 95, with the percentiles calculated based on the sub-cohort.

## Statistical analysis

### Probability weights

To derive population-based estimates from the case-cohort sample, we applied inverse probability weights in all analyses (except those limited to the subcohort). Cases (including those also ascertained through the subcohort) were assigned a weight of 1, and the non-cases were upweighted by (N_non-cases in the population_ /N_non-cases sampled_), as these observations represented a larger number of unobserved individuals in the source population. Because iPSYCH sampling was initially drawn from 1981-2005 births and then again from 1981-2008 births, we accounted for higher selection probability in those born before 2006. See **Table S3** for the weights calculations based on the case status and year of birth.

### Regression models

Each maternal diagnosis was analyzed for an association with child’s ASD PGS, first adjusting for the top 5 genetic PCs, child’s year of birth and legal sex (main models), and subsequently entering the additional demographic and socio-economic covariates (fully-adjusted models). For both crude and fully-adjusted models, we analyzed the association between each maternal diagnosis and child’s ASD PGS separately (single-diagnosis models), then included all maternal diagnoses in the same model to account for their co-occurrence (multi-diagnosis models).

To compare the associations between maternal diagnoses and ASD PGS vs. PGS for other mental health outcomes and height, we focused on single-diagnosis models only.

Throughout, we used linear regression models, implementing the svyglm function from the *survey*^*35*^ R package to incorporate the sampling weights, which account for the greater uncertainty around the observations with larger weights.

In additional analyses, we also explored whether maternal diagnosis is associated with differences in the tails of the child’s ASD PGS distribution, effects that may be missed when the focus is on the mean of the PGS score. We applied multinomial logistic regression using the *svyVGAM*^*36*^ package, categorizing ASD PGS as deciles, with decile 5 serving as the reference. The deciles were defined based on the subcohort and without the use of sampling weights.

To account for multiple testing, P-values were adjusted using the false discovery rate method^37^ to derive Q-values, using the function p.adjust. Statistical significance was defined as Q-value<0.05.

### Sensitivity analyses

In the sensitivity analyses, we first addressed differential availability of the diagnostic data based on maternal and child’s year of birth (as mothers born later in the calendar period had, on average, shorter follow-up periods, higher likelihood of receiving an ICD-10 (vs ICD8) diagnosis, and higher likelihood of receiving diagnoses in outpatient settings), by: (i) ensuring equal exposure time in all mothers, through narrowing the exposure window to the interval from 4 years prior to the birth of the child to 10 years after; (ii) ensuring availability of ICD-10 outpatient data for the entire exposure period, through restricting this analysis to children born after 1999; (iii) standardizing maternal ages for diagnosis ascertainment, through limiting the exposure window to maternal ages 18-45 (inpatient diagnoses only); we also re-run this analysis in the subset of mothers with available diagnostic information at those ages; (iv) verifying consistency of the effects across the different sources of the diagnosis (in-vs. outpatient) by limiting the exposure to diagnoses given in inpatient settings.

To verify the impact of the covariates in the fully adjusted (additional) analyses, we also (v) re-defined the healthcare utilization covariate to reflect diagnostic data from the 4 years prior to birth of the child to 8 years after, rather than lifetime, enabling us to standardize it across all mother-child dyads, and (vi) repeated the analyses controlling for family income (sum of maternal and paternal income), rather than only maternal income (in families with missing legal father, paternal income was set to 0).

Finally, we (vii) verified the impact of implementing sampling weights, by repeating the analysis in only the subcohort, using linear regression without weights (lm function from base R), and (viii) expanded the population to include both individuals of European and non-European ancestry.

## Results

### Sample

Our analytic sample consisted of 117,542 individuals, including 76,114 individuals ascertained based on the presence of a mental health diagnosis (see **Table S4** for the by-diagnosis breakdown) and 44,105 through the subcohort. Of those, 108,891 individuals were of European ancestry. **Table 1** presents the demographic characteristics of the European ancestry subset (see **Table S5** for the full sample demographics). Prevalences of maternal diagnoses tested for an association with child PGSs in the European ancestry individuals are presented in **Table S2**.

**Table 1.**
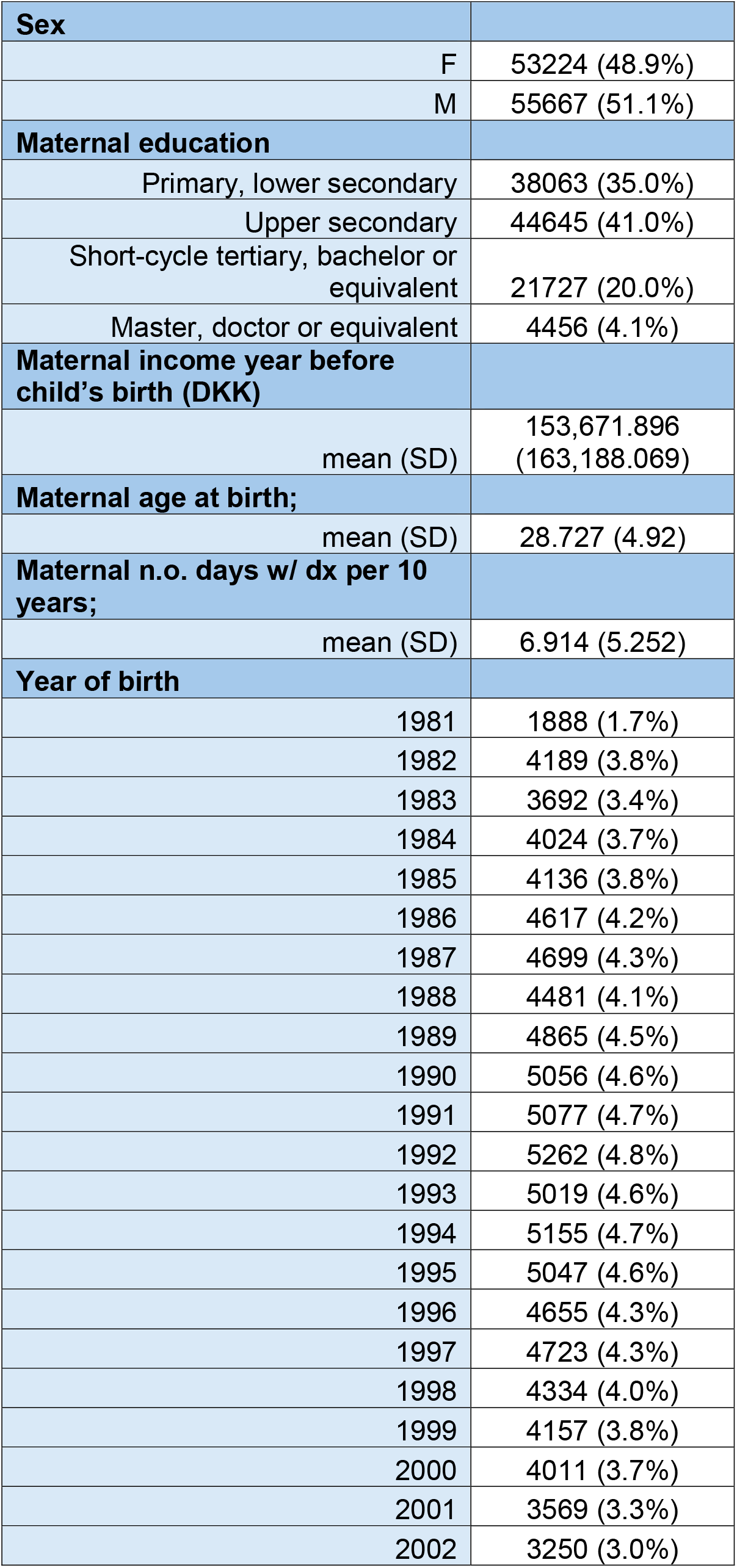
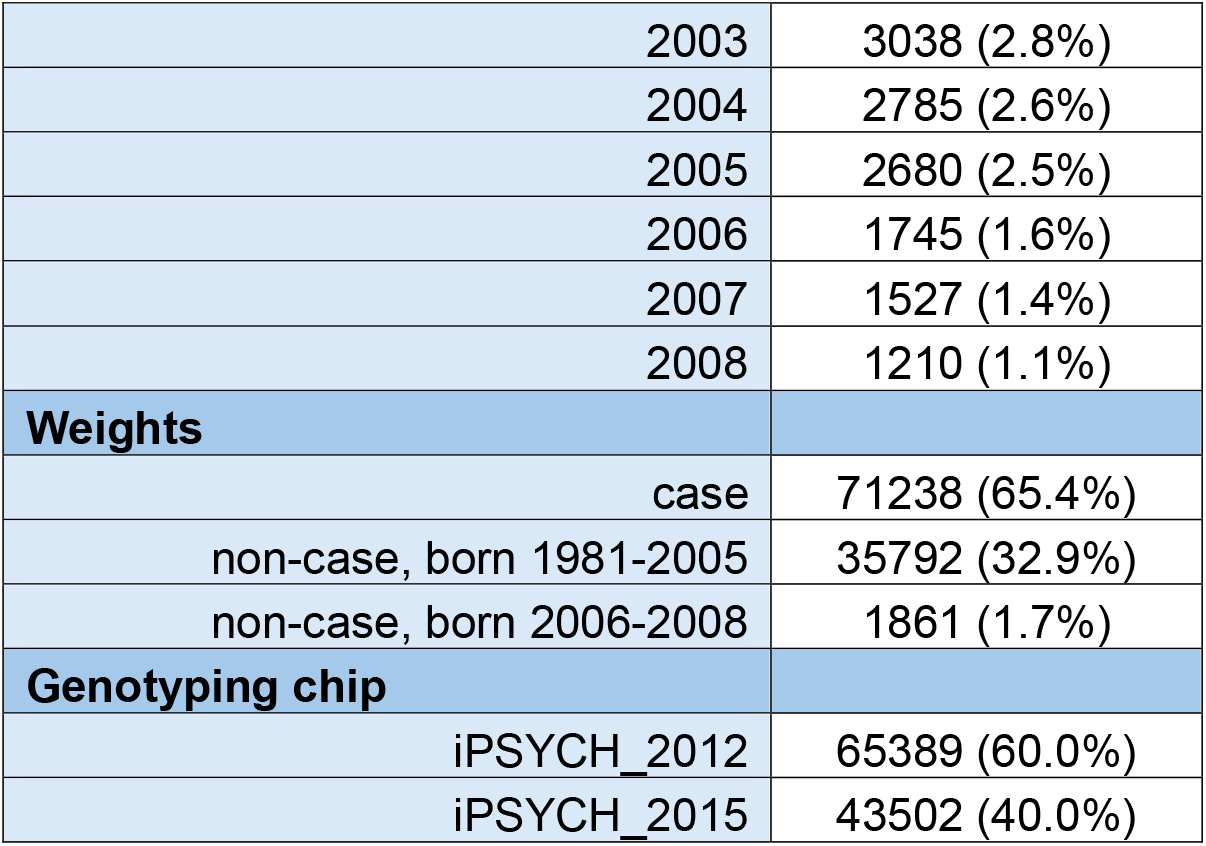
Demographic, sampling and genotyping characteristics of the analytical sample.

| Sex |  |
| --- | --- |
| F | 53224 (48.9%) |
| M | 55667 (51.1%) |
| Maternal education |  |
| Primary, lower secondary | 38063 (35.0%) |
| Upper secondary | 44645 (41.0%) |
| Short-cycle tertiary, bachelor or equivalent | 21727 (20.0%) |
| Master, doctor or equivalent | 4456 (4.1%) |
| Maternal income year before child's birth (DKK) |  |
| mean (SD) | 153,671.896<br>(163,188.069) |
| Maternal age at birth; |  |
| mean (SD) | 28.727 (4.92) |
| Maternal n.o. days w/ dx per 10 years; |  |
| mean (SD) | 6.914 (5.252) |
| Year of birth |  |
| 1981 | 1888 (1.7%) |
| 1982 | 4189 (3.8%) |
| 1983 | 3692 (3.4%) |
| 1984 | 4024 (3.7%) |
| 1985 | 4136 (3.8%) |
| 1986 | 4617 (4.2%) |
| 1987 | 4699 (4.3%) |
| 1988 | 4481 (4.1%) |
| 1989 | 4865 (4.5%) |
| 1990 | 5056 (4.6%) |
| 1991 | 5077 (4.7%) |
| 1992 | 5262 (4.8%) |
| 1993 | 5019 (4.6%) |
| 1994 | 5155 (4.7%) |
| 1995 | 5047 (4.6%) |
| 1996 | 4655 (4.3%) |
| 1997 | 4723 (4.3%) |
| 1998 | 4334 (4.0%) |
| 1999 | 4157 (3.8%) |
| 2000 | 4011 (3.7%) |
| 2001 | 3569 (3.3%) |
| 2002 | 3250 (3.0%) |

|  |  |  |
| --- | --- | --- |
|  | 2003 | 3038 (2.8%) |
|  | 2004 | 2785 (2.6%) |
|  | 2005 | 2680 (2.5%) |
|  | 2006 | 1745 (1.6%) |
|  | 2007 | 1527 (1.4%) |
|  | 2008 | 1210 (1.1%) |
| **Weights** |  |  |
|  | case | 71238 (65.4%) |
|  | non-case, born 1981-2005 | 35792 (32.9%) |
|  | non-case, born 2006-2008 | 1861 (1.7%) |
| **Genotyping chip** |  |  |
|  | iPSYCH_2012 | 65389 (60.0%) |
|  | iPSYCH_2015 | 43502 (40.0%) |

### Association between maternal health and ASD PGS

In the main models, several maternal diagnoses were nominally significantly associated with ASD PGS, including *fetal distress* (ICD10 O68; MD (mean difference)= 0.03 [0.01, 0.06], P=0.018); *injury of eye and orbit* (ICD10 S05; MD=0.07 [0.02, 0.12], P=0.007); *obesity* (ICD10 E66; MD=0.04 [0.01, 0.08], P=0.012); and *asthma* (ICD10 J45; MD=0.06 [0.02, 0.11], P=0.006). However, none of these associations remained significant after the multiple testing correction (Q<0.05; **Table S6; Figure 1**).

**Figure 1:**
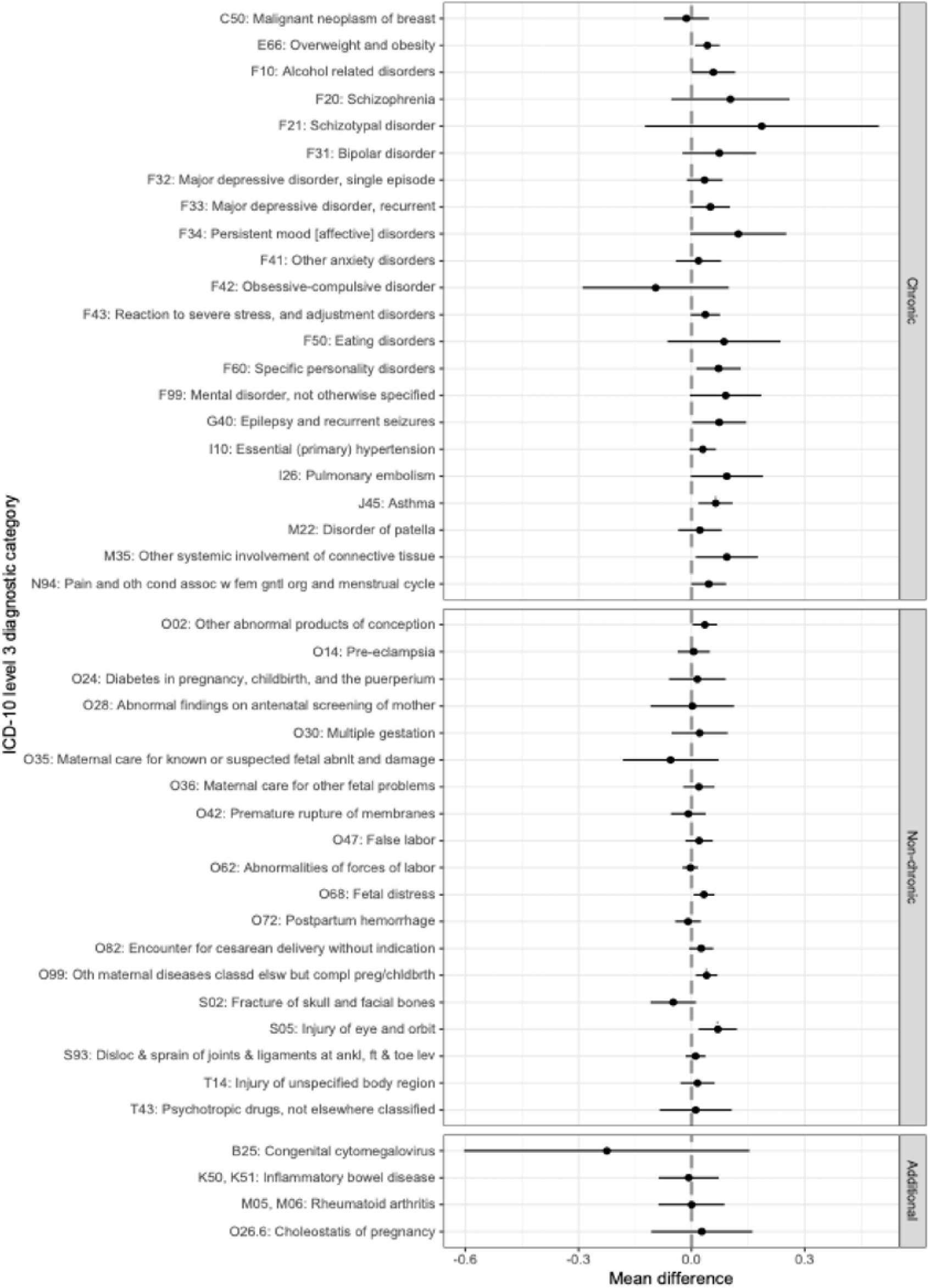
Results from weighted linear regression models comparing ASD PGS in children who were exposed/unexposed to maternal conditions listed on the y-axis. Point estimates are derived from single diagnosis models and reflect the mean difference in ASD PGS between the groups with its 95% confidence interval. All models were adjusted for genotyping round/chip, principal component 1-5, child’s sex and year of birth.

These results were stable in the additional models, where we included maternal demographic and socio-economic factors as covariates (**Table S6**), as well as across the sensitivity analyses (see below). We also observed consistency of the results in the multi-diagnosis model, except for the attenuation of the estimates for maternal psychiatric diagnoses after this adjustment for other maternal conditions (**Table S6**).

### Association between maternal health and other PGS

The associations between maternal diagnoses and PGS of other neuropsychiatric conditions were in a consistent direction as those for ASD PGS, but typically stronger and more often statistically significant (Q < 0.05; Error! Reference source not found.Error! Reference source not found.; **Table S9**). For many of the maternal diagnoses, we observed the strongest associations (largest effect sizes) with children’s MDD, SCZ and ADHD PGSs. The strongest positive associations were between the child’s SCZ PGS and maternal diagnoses of schizophrenia and schizotypal disorder.

Similar to ASD PGS, these associations were largely stable in the model adjusted for the additional covariates. The most pronounced differences between the crude and fully adjusted model were recorded for ADHD PGS (**Table S9**), where none of the associations that were statistically significant in the main model remained so after additional adjustment for these covariates.

Height PGS, used as a negative control outcome, was statistically significantly associated with three maternal diagnoses: *fetal distress* (ICD10 O68), *postpartum hemorrhage* (ICD10 O72), and *specific personality disorder* (ICD10 F60; **Figure 2; Table S9**).

**Figure 2:**
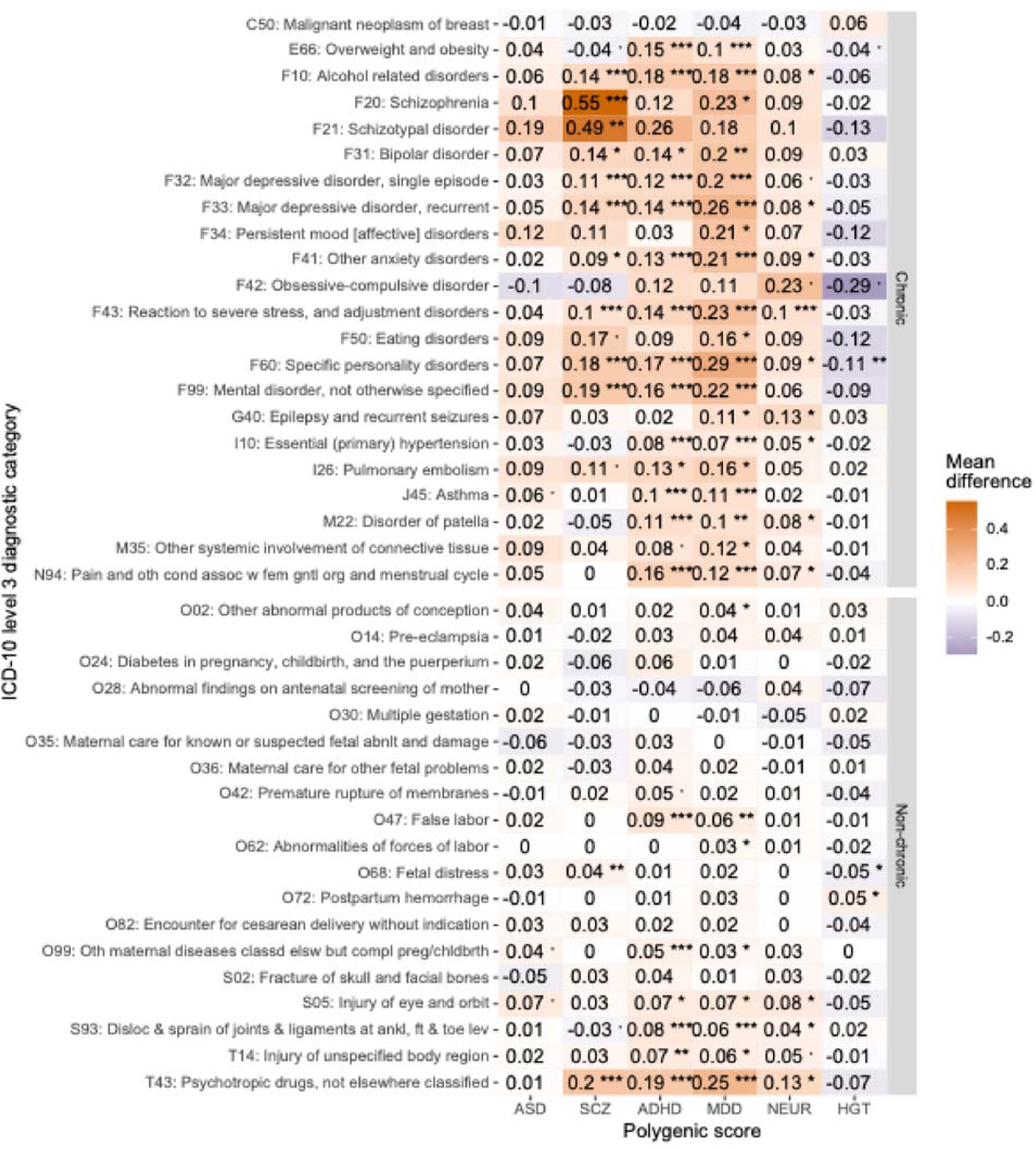
Results from weighted linear regression models comparing PGS in children who were exposed/unexposed to maternal conditions listed on the y-axis. (ASD=autism spectrum disorder; SCZ=schizophrenia; ADHD=attention-deficit/hyperactivity disorder; NEUR=neuroticism; HGT=height). Point estimates are derived from single diagnosis models and reflect the mean difference in the respective PGS between the groups. All models were adjusted for genotyping round/chip, principal component 1-5, child’s sex and year of birth. Mean difference is displayed together with stars indicating q-value smaller than: 0.1, * 0.05, ** 0.01, ***0.001.

### PGS differences in the distribution of tails in relation to maternal health

For several maternal diagnoses, the multinomial regression results based on the tails of the ASD PGS distribution behaved as expected given the associations of maternal diagnoses with the ASD PGS mean: i.e., when children of exposed mothers had a higher *mean* ASD PGS in the main analyses, we observed that they were less likely to be in the bottom PGS decile, and more likely to be in the top decile, compared to their likelihood of being in the middle (5^th^) decile (e.g., *schizotypal disorder*, ICD10 F21; OR_1v5_=0.57 [0.14, 2.23], P=0.415; OR_10v5_=2.21 [0.79, 6.15], P=0.129), although this pattern was not consistent across all deciles (see **Table S10, Figure 3**).

**Figure 3:**
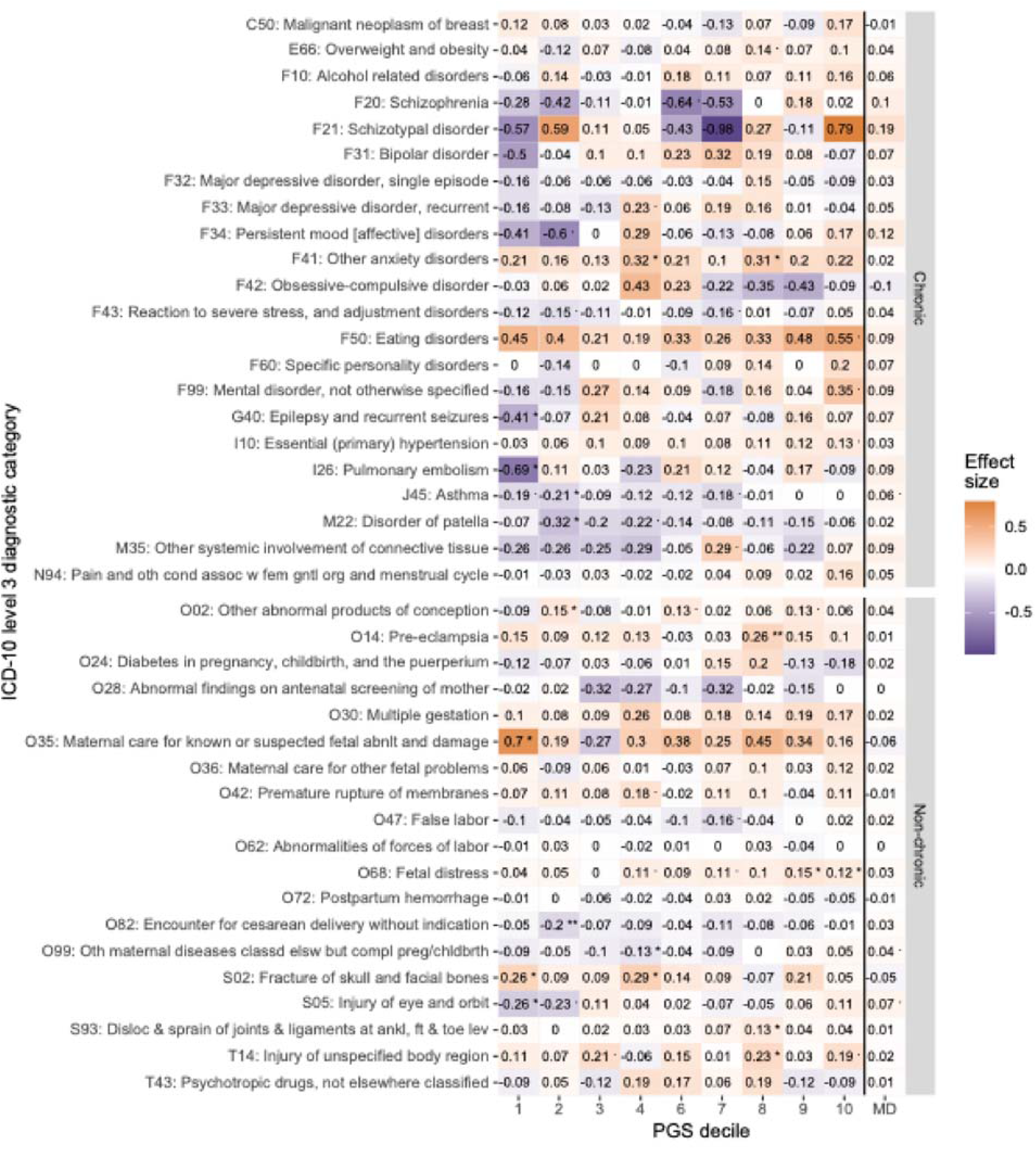
Results from weighted multinomial logistic regression models (log(OR), 1-4 and 6-10 on the x-axis) and weighted linear regression models (MD on the x-axis) comparing ASD PGS in children who were exposed/unexposed to maternal conditions listed on the y-axis. Point estimates are derived from single diagnosis models and reflect the mean difference in ASD PGS between the groups (MD), and the log odds ratios of being in the different ASD PGS deciles compared to decile 5, in exposed vs. unexposed children. All models were adjusted for genotyping round/chip, principal component 1-5, and the child’s sex and year of birth. On the figure, effect size is displayed together with stars indicating p-value smaller than: 0.1, * 0.05, ** 0.01, ***0.001.

However, we also observed associations where the differences in ASD PGS means were driven by the exposed children being less likely have low ASD PGS (i.e., be in the bottom ASD PGS decile), including e.g., *epilepsy* (ICD-10 G40; OR_1v5_=0.66 [0.45, 0.97], P=0.035; OR_10v5_=1.07 [0.76, 1.52], P=0.69) and *pulmonary embolism* (ICD-10 I26; OR_1v5_= 0.50 [0.28, 0.89], P=0.017; OR_10v5_= 0.91 [0.55, 1.52], P=0.723). See **Table S10**.

### Sensitivity and additional analyses

Our results remained largely consistent across a series of sensitivity analyses, including (a) restriction of the sample to the subcohort (**Table S11**), (b) standardizing the exposure windows to between 4 years pre- and 10 years post-childbirth (**Table S12**), (c) re-defining the healthcare utilization measure (**Table S13**); and (d) including paternal income in “household income” and controlling for it in the regression models (**Table S14**). While most effects remained consistent after (e) inclusion of inpatient diagnoses only, we observed the largest differences between the main and this sensitivity analysis for two of the diagnoses in this sensitivity analysis: obsessive-compulsive disorder (OCD) and eating disorders, with, respectively, higher and lower effect sizes after restriction to inpatient diagnoses (**Table S15**). These effects on eating disorders were further magnified when we limited the analytical sample to mothers aged 18-45 (**Table S16**). Finally, all results were consistent between European-only and the entire (multi-ancestry) iPSYCH sample (**Table S8**).

## Discussion

We have observed that a lifetime maternal diagnosis of multiple conditions is associated with higher polygenic propensities to neuropsychiatric conditions in offspring. While the maternal conditions associated with child neuropsychiatric PGSs spanned a broad medical spectrum (e.g., obstetric and injury codes), the majority of the associations were observed for the maternal mental health diagnoses — consistent with the earlier studies demonstrating considerable genetic correlations across psychiatric conditions^27^. Children’s non-autism PGSs, e.g., those for MDD or schizophrenia, were typically more strongly associated with maternal health than their ASD PGS, with these effects also more likely to be statistically significant.

The overarching goal of our study was to investigate whether the observational associations between maternal health and offspring autism^9^ are, at least in part, attributable to polygenic effects captured by PGS. Inclusion of a broader range of neuropsychiatric PGS was motivated by strong genetic correlations between those diagnoses and autism^26,27^, and the availability of more powerful GWASs available for those conditions – due to both larger case-control samples in those studies, and the (likely) lower fraction of cases with a monogenic etiology (who contribute less to the polygenic signal^38,39^). While we cannot rule out that the stronger results for non-autism PGSs reflect unique etiological pathways between maternal health and child genetic liabilities for those conditions, a more parsimonious interpretation is that the autism and non-autism PGSs capture similar familial signal, but the latter PGSs are better powered to detect it.

Our results indicate that the observational associations between maternal health and child neuropsychiatric outcomes arise, at least in part, due to genetic effects. However, as the magnitude of the associations between maternal conditions and child PGSs in our study was overall low, they did not, on their own, fully account for the epidemiological observations^9^. This could indicate that these observational associations are underlain by additional mechanisms, including e.g., other familial, non-genetic influences (for example, family’s socioeconomic status, which can affect the likelihood of both certain maternal diagnoses and autism, and/or contribute to an earlier diagnosis of the latter). Alternatively, our results could suggest that the “genetic effects” captured in our study do not fully capture the range of genetic mechanisms underlying the associations between maternal health and autism. This could occur when (i) the polygenic effects contributing to these associations are not fully captured by the PGS; (ii) if the observational associations arise, at least in part, due to the effects of rare variants, which are not included in the majority of the PGS models, but which have a substantial role in autism etiology^40,41^, including those shared by the family members^42,43^; and/or (iii) the relevant genetic effects go beyond the direct mother-to-child transmission, instead being influenced also by e.g., indirect mechanisms^18,44^, which do not rely on allelic transmission. As the PGSs in our study were constructed based on child, rather than maternal genotype data, the estimates of such indirect (maternal) effects operating by influencing prenatal environment would be attenuated in the offspring generation. Therefore, while the observational associations between maternal health and autism cannot be fully attributed to genetic mechanisms based on our study, the plausibility of all of i-iii reinforces that such epidemiological observations need to continue to be carefully scrutinized for potential genetic confounding.

Our results were robust to a series of sensitivity analyses, including truncating the exposure time to standardize it across maternal birth years, performing the analysis in the subcohort sample only, or including the non-European ancestry participants. Restricting the exposures to maternal diagnoses given only in the inpatient settings resulted in reversal of the direction of the effects for maternal OCD and eating disorders, but had minimal impact on the remaining associations. Lastly, accounting for maternal comorbidity by entering all maternal diagnoses as joint predictors of child’s ASD PGS, had the strongest effects on the associations between ASD PGS and maternal mental health conditions. This is in line with our epidemiological observations^9^, and the evidence of extensive comorbidity between mental health and other conditions, exceeding the one observed between other pairs of somatic conditions^45^.

While the effects of maternal health on height PGS were mostly not statistically significant, we did observe an association between fetal distress and postpartum hemorrhage with, respectively, lower and higher height PGS. The differential direction of these effects suggests that these results are unlikely to reflect a systematic inflation/deflation of our results. Instead, the literature suggests that height may be an imperfect negative control due to its associations with reproductive^46^ traits.

Examining the full PGS distribution using multinomial models suggested potential differences not captured by comparing the mean PGSs, including e.g., children born to mothers with epilepsy or pulmonary embolism having lower odds of being in the lowest decile of the ASD-PGS distribution.

The strengths of the study include a broad ascertainment of maternal conditions observationally associated with autism, offering a comprehensive overview of the role of common genetic variation in those associations. Use of the population-based iPSYCH sample enabled ascertainment of maternal and child diagnoses through national registers – enhancing the study’s reliability and allowing us to calculate population-based estimates based on known selection weights. Lastly, using meta-PGS enabled us to augment PGSs trained on external data by leveraging our large-scale, internal iPSYCH resources, without overfitting.

The limitations include the lack of genotype data from broader pedigrees, which precluded differentiating direct and indirect effects, as well as estimating the impact of assortative mating^47–49^ — whereby females with certain genetic liabilities are more likely to have partners with similar genetic profiles, with consequences on the child PGS^50^. Additionally, PGS, despite their high utility in summarizing genetic liabilities across a variety of traits, still explain lower trait variance than could be expected from SNP-based heritability^51,52^. This is further exacerbated by the involvement of rare variants in autism^41^, resulting in a likely under-estimation of genetic effects in our study. Additionally, our cohort spanned a wide range of maternal and childbirth years, introducing heterogeneity into clinical diagnoses (ICD-8 and ICD-10) and their ascertainment (nationwide availability of outpatient diagnoses). To account for this, we controlled for childbirth year and maternal age, and implemented a series of sensitivity analyses, all of which indicated that the impact of this long ascertainment period was very limited.

In conclusion, we observed that maternal conditions in pregnancy – predominantly psychiatric ones, but also obstetric conditions and injury - are associated with the child’s PGSs for neuropsychiatric diagnoses. These effects were stronger for non-ASD PGS, although these differences could reflect the power of the underlying GWAS and the genetic architecture of the respective conditions. As even the strongest associations are unlikely to account for the epidemiological results, these observational associations are likely to arise also due to non-genetic effects, and/or genetic mechanisms not captured by the current PGSs.

## Data availability

The Danish Scientific Ethical Committee system, as well as all relevant register authorities, including the Danish Data Protection Agency, Statistics Denmark and the Danish Health Data Authority, approved access to the data under strict conditions regarding access and data export. Under these conditions, there are no provisions for exporting individual-level data, all or in part, to another institution in or outside of Denmark. The minimum datasets, including all summary statistics of the measures of associations used to draw the study conclusions, are presented in the supplementary material.

## Code availability

Analytical code is available upon request

## Acknowledgements

This work was supported by grants from the National Institute of Mental Health (R01MH124817, T32-MH122394), the Eunice Kennedy Shriver National Institute of Child Health and Human Development (R01HD098883), National Institute of Neurological Disorders and Stroke (R01NS131433), and the Lundbeck Foundation (iPSYCH, grant nos. R102-A9118 and R155-2014-1724).

## Supplemental Figures

**Figure S1.**
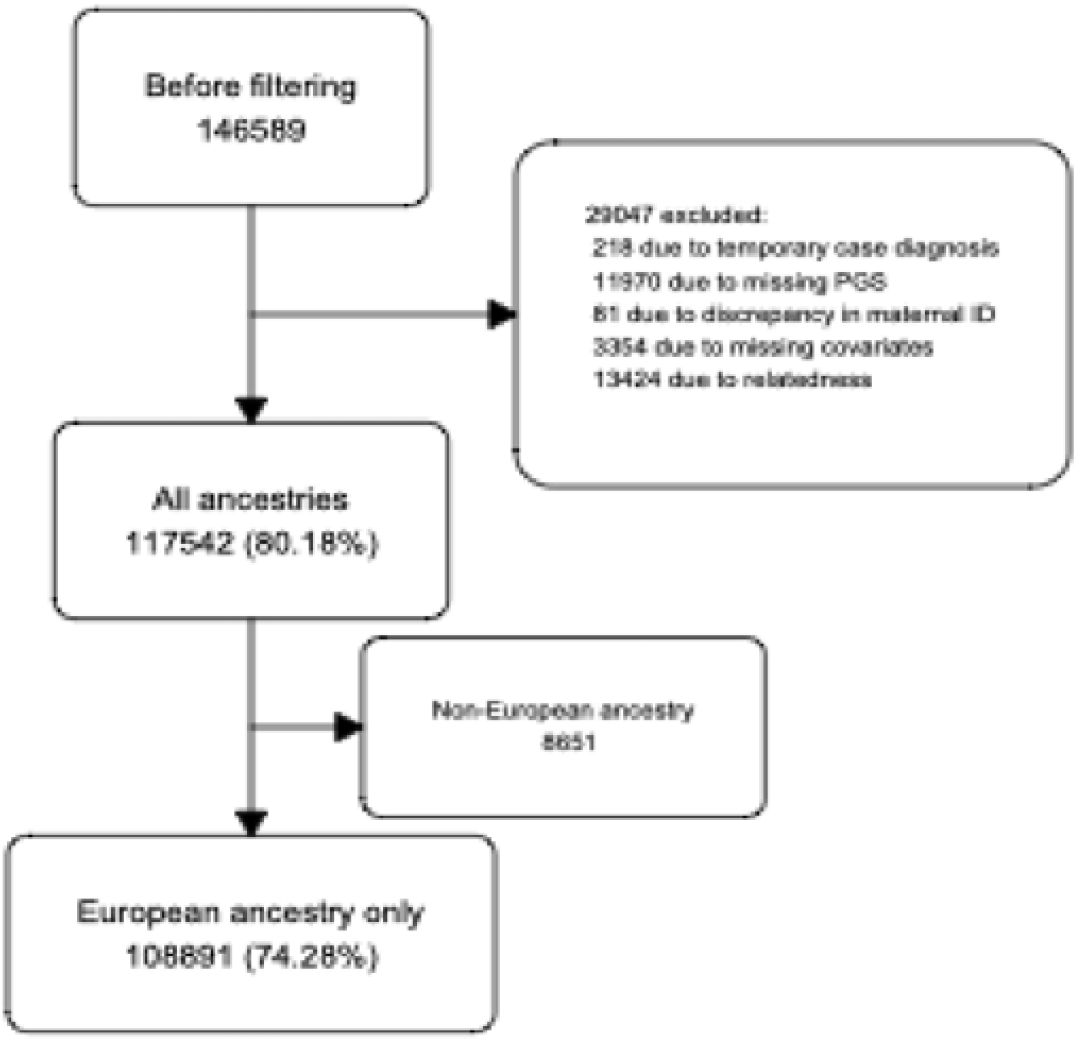
Flowchart of sample processing and quality control.

